# Inverse planning for radiofrequency ablation in cancer therapy using multiple damage models

**DOI:** 10.1101/2021.06.07.21258239

**Authors:** Shefali Kulkarni-Thaker, Dionne Aleman, Aaron Fenster

## Abstract

Radiofrequency ablation (RFA) offers localized and minimally invasive treatment of small-to-medium sized inoperable tumors. In RFA, tissue is ablated with high temperatures obtained from electrodes (needles) inserted percutaneously or via an open surgery into the target. RFA treatments are generally not planned in a systematic way, and do not account for nearby organs-at-risk (OARs), potentially leading to sub-optimal treatments and inconsistent treatment quality. We therefore develop a mathematical framework to design RFA treatment plans that provide complete ablation while minimizing healthy tissue damage. Borrowing techniques from radiosurgery inverse planning, we design a two-stage approach where we first identify needle positions and orientations, called needle orientation optimization, and then compute the treatment time for optimal thermal dose delivery, called thermal dose optimization. Several different damage models are used to determine both target and OAR damage. We present numerical results on three clinical case studies. Our findings indicate a need for high source voltage for short tip length (conducting portion of the needle) or fewer needles, and low source voltage for long tip length or more needles to achieve full coverage. Further, more needles yields a larger ablation volume and consequently more OAR damage. Finally, the choice of damage model impacts the source voltage, tip length, and needle quantity.

## 1 Introduction

Hepatocellular carcinoma is the fifth-most common diagnosed malignancy, and the third-most frequent cause of cancer-related deaths world-wide (Bhardwaj et al, 2010; Rempp et al, 2011; WHO, 2012). Although surgical resection is the preferred treatment choice for liver cancer, up to 80% of these patients cannot be operated on due to tumor location or existing co-morbidities (Bhardwaj et al, 2010; O’Rourke et al, 2007; Pearson et al, 1999). Focal ablation (removal of tissue with extreme temperatures), a localized, minimally invasive treatment option for small-to medium-sized (Rempp et al, 2011) tumors with fewer side effects than surgery, radiation, or chemotherapy, is then the treatment option for such patients. Ablation has been used to treat several cancer sites including liver, abdomen, renal, lung, and prostate (Dupuy et al, 2000; Eggener et al, 2007; Livraghi et al, 2003; Pavlovich et al, 2002). It has few side effects, short recovery times, out-patient delivery, and minimal organs-at-risk (OARs) damage due to localized treatment (Goldberg et al, 2000). There are several ablative therapies including microwave ablation (MWA), radiofrequency ablation (RFA), high intensity focused ultrasound (HIFU), laser ablation, and cryoablation. We present a systematic approach to deliver RFA treatments by quantifying tissue damage using several different models. This framework is applicable to other ablation modalities.

In RFA, a needle is inserted into the target (tumor or lesion) percutaneously or via open surgery, and high frequency alternating current is passed through the needle. Heat is generated due to tissue resistance and the target achieves necrosis when exposed to high temperatures for an adequate amount of time. Despite its appealing characteristics, RFA has the potential for incomplete ablation, and therefore a high local recurrence (regrowth) (Bhardwaj et al, 2010), when the tumor is close to large blood vessels, the needle is incorrectly placed, or insufficient heat is delivered. Needles are typically guided into position with the assistance of ultrasound imaging (Neshat et al, 2013), with the patient optionally sedated, though patient cooperation can be useful in some situations, particularly liver and lung ablation (Piccioni et al, 2019).

In inverse planning, the target and OARs are divided into unit grids called voxels (“volume pixels”) and optimal doses are sought for these structures. This methodology has been previously used successfully to plan cancer treatments using radiosurgery (Ferris and Shepard, 2000; Ferris et al, 2002; Ghobadi et al, 2012, 2013) and intensity modulated radiation therapy (Romeijn and Dempsey, 2008). Unfortunately, unlike radiation treatments, where dose delivered from several beams is additive, heat delivered from multiple needles is not directly additive and must be calculated using Pennes’ bioheat transfer equation (BHTE), a partial differential equation (PDE) (Wissler, 1998). Further, BHTE does not consider the amount of time a voxel is exposed to a temperature. Alternate thermal damage models, e.g., the Arrhenius thermal damage model (ATDM) (Henriques, 1947; Henriques and Moritz, 1947; Moritz, 1947; Moritz and Henriques, 1947) and cumulative equivalent minutes at 43^°^C (CEM_43_) (Sapareto and Dewey, 1984), use a voxel’s temperature history, obtained from BHTE, during the course of treatment to determine tissue thermal damage. These models, although nonlinear, are additive across multiple needles. Thus, the development of inverse plans for ablation is mathematically challenging due to the inherent nonlinear nature of thermal damage. Because RFA operates at temperaturs ≥60^°^C, CEM_43_ is not an appropriate model for this ablation therapy.

Existing work on RFA inverse planning can be categorized into exact and inexact methods. Inexact methods approximate the ablated region to a sphere or an ellipse of a known fixed size based on the needle used (Butz et al, 2000; Mundeleer et al, 2009; Villard et al, 2005; Zhang et al, 2007). A voxel within the ablated sphere or ellipse is considered destroyed. Thus, there is no actual dose computation and the needle is positioned by unconstrained optimization models solved using Powell’s (Butz et al, 2000; Villard et al, 2005; Zhang et al, 2007) or Nelder-Mead (Downhill Simplex) (Mundeleer et al, 2009; Villard et al, 2005) algorithms. The objectives of these models is typically to maximize the difference in unablated target and OAR volumes for single (Butz et al, 2000; Villard et al, 2005; Zhang et al, 2007) or multiple RFA applicators (Mundeleer et al, 2009). Although these methods are fast and the assumption of knowing the ablation radii a priori is plausible, they do not consider the presence of cooling effects like blood perfusion and therefore may result in incomplete ablation.

Exact methods (Altrogge et al, 2007; Chen et al, 2006, 2009; Haase et al, 2012) compute the thermal dose received by a voxel at each time step using BHTE where the energy absorbed due to the radiofrequency electric source, referred to as the specific absorption rate (SAR), is obtained using the Laplacian equation, an electrostatic PDE. Thus, there is no prior assumption on ablation radii. The decision variables in exact models are the position and the orientation of the needle with fixed treatment time, and hence, needle positioning and thermal dose optimization are simultaneously performed. The resulting models are nonlinear, constrained by a system of PDEs describing the electric potential of the applicator and steady-state BHTE with (Haase et al, 2012) or without (Altrogge et al, 2007) consideration of risk structures (e.g., ribs), and are solved using gradient-based optimization methods (Altrogge et al, 2007; Haase et al, 2012). Models that use the Arrhenius-based thermal damage model to minimize the survival fraction of the target using steepest descent (Chen et al, 2006, 2009) have better computational tractability. Since needle positioning and thermal dose computation happen simultaneously, these models require computation of a PDE, a computationally intensive task, at every new needle position and orientation. Further, these methods are tailored to RFA-specific PDEs (electrostatic field) and therefore cannot be immediately applied to MWA (electromagnetic field) or HIFU (acoustic field).

We approach the RFA inverse planning problem in two stages, incorporating ideas from exact as well as inexact methods. In the first stage, called needle orientation optimization (NOO), we use geometric shape approximations to compute needle position and orientation for single needle and multiple independent single needle scenarios, where clustered needles with relative fixed positions are treated as a single needle. In the second stage, we use the NOO solution to optimize the duration and voltage of the needles using BHTE; this stage is called thermal dose optimization (TDO), and several common thermal damage models are explored.

Using the solution from NOO, we enumerate solutions for different damage models by pre-computing them for a fixed treatment time for different source voltages and needle types. We then perform a look- up to identify minimum source voltage and treatment time based on full target coverage. Using this large amount of information, we are able to perform several different analyses, including change in target and OAR coverage volumes against increasing source voltages and needle types. Finally, we identify the best needle configuration that gives full target coverage and the least OAR damage.

### Algorithm 1

Focal ablation framework

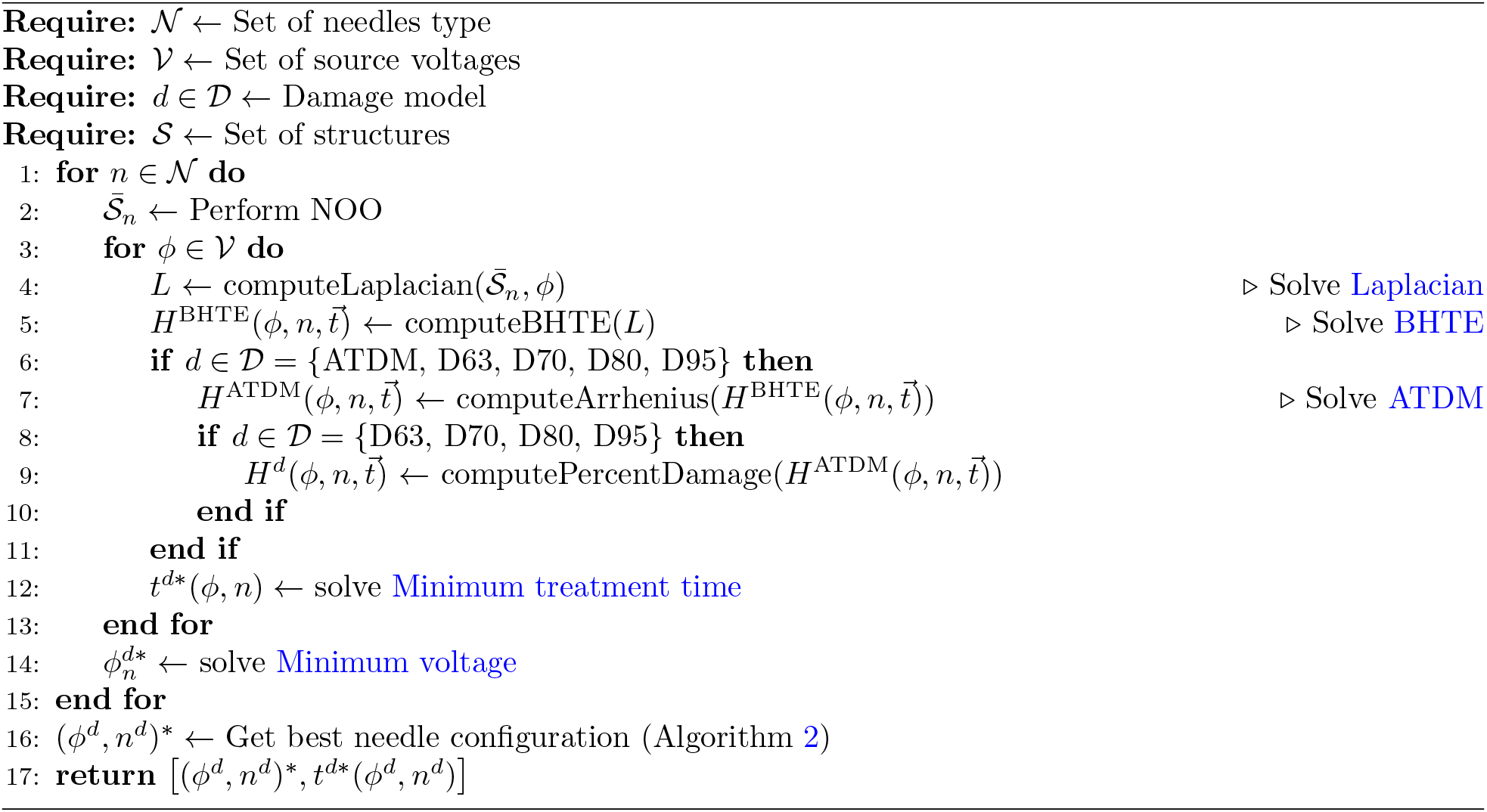

Our methodology has several benefits over current strategies, both in terms of computational methodology and clinical practicality. Methodologically, we eliminate the need to compute BHTE at each iteration, thereby saving several hours of computation time, and we eliminate geometric assumptions from NOO in TDO where we compute the actual thermal dose delivered. By pre-computing damage models, we again save computation time and are able to analyze target damage at different source voltages. With respect to clinical practicality, we are able to determine OAR damage, which we exploit to determine the best needle configuration for treatment. The detailed analysis enables the decision maker to obtain a complete treatment plan for the preferred damage model. Finally, we can also extend our framework to other focal ablation methods like MWA by solving a different set of PDEs instead of BHTE.

The remainder of this paper is organized as follows. Section 2 presents the methodology, including NOO and TDO. Numerical results and discussion are provided in Section 3 and 4, respectively.

## 2 Methodology

Let 𝒯 be the set of target structures. Voxels not identified as target form the OAR set, ℋ. Further, let 𝒩, *𝒱*, and 𝒟 be the set of needles, source voltages, and damage models, respectively. For *n* ∈𝒩, we perform NOO for a target in 𝒯. Using the computed needle position and orientation, we determine thermal damage to the target for each *ϕ* ∈*𝒱*. Finally, we identify the best needle configuration ((*ϕ, n*) ∈𝒱× 𝒩) and treatment time. The complete framework is presented in Algorithm 1.

### 2.1 Needle orientation optimization

We explore NOO for three kinds of needle configurations: (i) single needle, (ii) clustered needles, and (iii) 2-3 independent needles (called multiple needles) (Medtronic, 2016). A clustered needle is a single device with three parallel tines (needles) that operate simultaneously. Multiple needles are multiple single needle devices that can be inserted randomly or in parallel. They can either be operated simultaneously or individually to ablate a larger volume or several smaller volumes, though simultaneous operation is clinically more common for individual target volumes as heat damage (scarring) from previous needles in sequential ablation results in unpredictable ablation to remaining untreated cells. In practice, a single needle configuration is used according to specific equipment availability and the preference of the interventional radiologist performing the procedure, and we therefore present models for each configuration, allowing interventional radiologists to chose the model that corresponds to their clinical preferences. We assume that the needles are inserted in a random order and are operated simultaneously.

Vendor specifications indicate an ellipsoidal (Figure 1(a)) and spherical (Figure 1(b)) shape of thermal lesions for single and clustered RFA electrodes, respectively (Medtronic, 2016).

**Figure 1:**
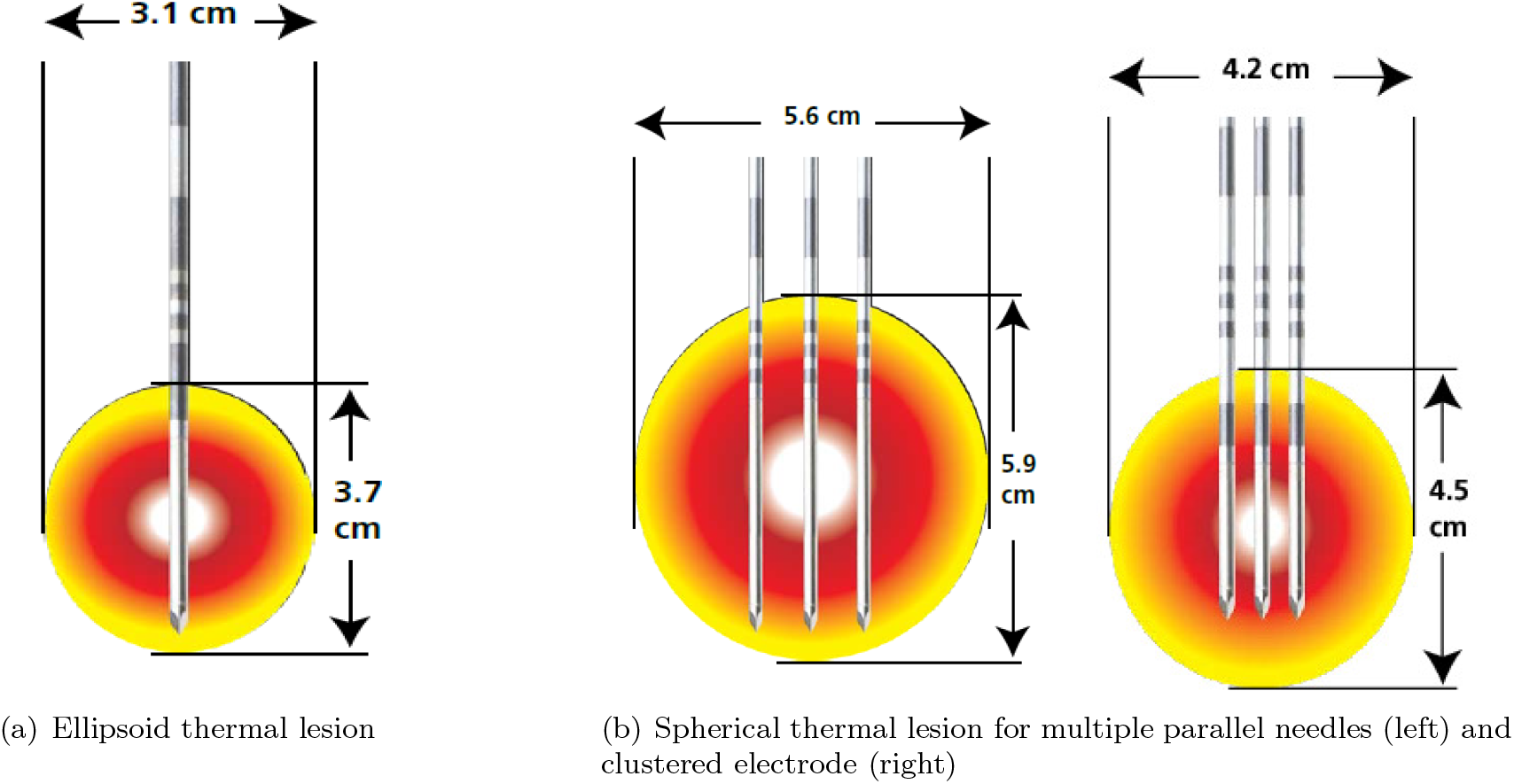
Cool-tip™ RF Ablation System E Series electrodes (Medtronic, 2016)

The NOO problem can therefore be represented as fitting a smallest ellipse (minimum volume covering ellipse, MVCE) or sphere (minimum volume covering sphere, MVCS) around the target. The central axis of the fitted ellipse or sphere is the orientation of the needle. We formulate the MVCE and MVCS optimization models as follows. Let 𝒯 ′ ⊂𝒯 be the set of target boundary voxels, 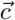 be the center of the needle, 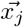 be the coordinate position for voxel *j* ∈*𝒯*.

Using a fixed space separator, multiple electrodes placed parallely and operated simultaneously assume a spherical lesion and can be treated as a clustered electrode (Figure 1(b)). However, when needles are inserted randomly, geometric shapes are unclear. Therefore, for the purpose of NOO, in the case of *k* multiple needles, we treat each needle as though it were operated independently and divide the tumor into *k* clusters where each cluster corresponds to single needle coverage. A similar approach was used by Chen et al (2009). The orientations are determined by MVCE and the entire process is referred to as NOO-Kmeans. The independent needle operation assumption is later relaxed in TDO.

#### 2.1.1 Single and clustered needle placement

From basic algebra, the equation of an ellipse in *m* dimensions with center (*c*_1_, …, *c*_*m*_) and radii (*a*_1_, …, *a*_*m*_) is given by

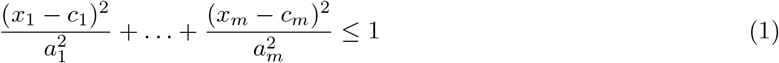

where *x*_1_, …, *x*_*m*_ are coordinates of target voxels inside the closed ellipse. Using matrix notation, we can rewrite Equation 1 as a set of points, *ξ*:

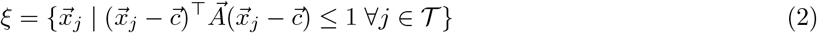

where 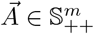, a set of *m* × *m* symmetric positive definite matrices, is full rank, and *m* is the dimensionality of the matrix 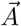, which in our case is 3 since the target is 3D. The eigenvalue decomposition of matrix 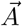 is given by 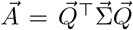, where 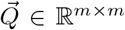 is an orthonormal matrix representing the eigenvectors of 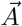 and 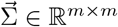 is a diagonal matrix whose entries (*λ*_1_, …, *λ*_*m*_) represent the eigenvalues of 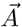. From the Principle Axis Theorem, the square root of the inverse of the eigenvalues represent the length of each semi-axis of the ellipse, i.e., 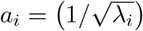, while the eigenvectors, i.e., columns of 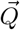, represent its orientation. The volume of an ellipse is therefore proportional to 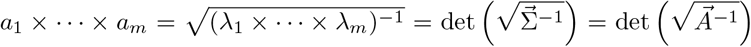, where det() is the determinant.

For any 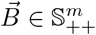, the determinant is given by

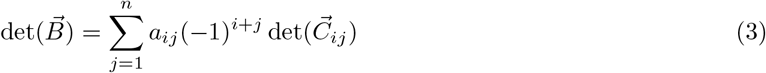

where 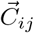 is the minor matrix obtained by dropping row *i* and column *j* from matrix 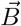. The determinant is a high-degree polynomial and therefore we perform Cholesky decomposition on 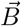 and use the simplified convex log det() function. We decompose 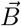 into lower triangular matrices, 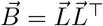 where 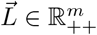. Thus, 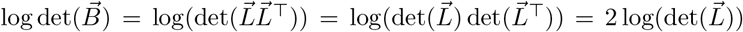. Since 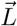 is a lower triangular matrix, its determinant is simply the product of its diagonal entries, i.e., 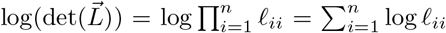.

Thus, 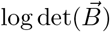 is convex.

Now, we formulate a convex mathematical model to find the minimum volume covering ellipsoid (MVCE) covering a set of finite points using a log det() function (Boyd and Vandenberghe, 1996):

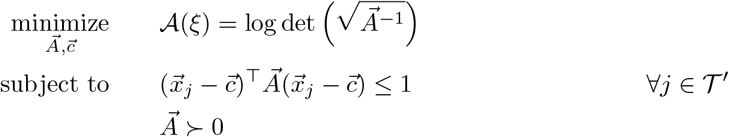

where ≻ enforces positive definiteness on 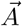. This model has a convex objective function, but a non-convex constraint. However, the constraint can be reformulated as convex by defining 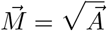 and 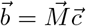:

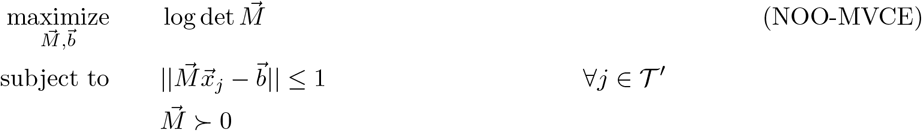

This problem is a maximization of a concave function with convex constraints and is thus a convex optimization problem, which can be solved to optimality. From the global optimal solution, 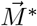 and 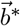, we can obtain needle orientation and position by 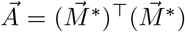 and 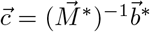, respectively. The eigenvalue decomposition of 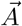 will give the orientation and stretch of the ellipse as described before. The eigenvector corresponding to the smallest eigenvalue represents the longest semi-axis of the ellipse and hence gives the orientation of the needle. The center 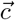 corresponds to the center of the conducting part of the needle.

We can solve NOO-MVCE to optimality since it is a maximization of a concave function with convex constraints and is therefore convex. We can obtain the needle position by 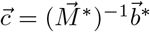 and orientation by eigenvalue decomposition of 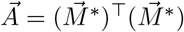.

We treat a clustered needle applicator (Figure 1(b)) as a single needle, and identify smallest sphere covering all the voxels by formulating a convex MVCS optimization model with an affine objective and a second-order conic constraint (Boyd and Vandenberghe, 2004):

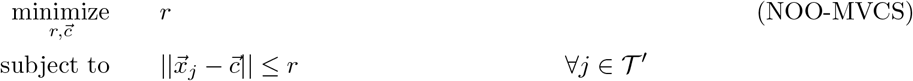

where *r* and 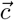 are the radius and center of the sphere, respectively. The tines in the clustered applicator are fixed, parallel, and equidistant. Therefore, the center of the fitted sphere corresponds to the barycenter or centroid of the equilateral triangle formed by the centers of the conducting tips of each tine in the cluster. We choose the cluster orientation along the diameter that maximizes needle-tumor contact. If the conducting tines overlaps non-target voxels, then we rotate the cluster in increments of 5^°^ until we find a better needle- tumor contact. We note that the equilateral triangle has a rotational symmetry of 120^°^, which means the triangle (or the needles of the cluster) maps onto itself after 120^°^ rotation. Thus, we explore only 24 cluster rotations in a given direction (Figure 2).

**Figure 2:**
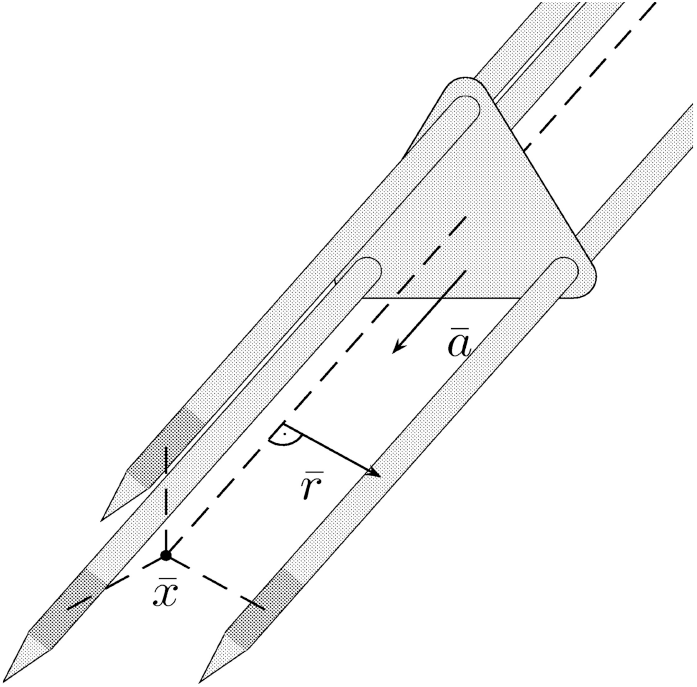
Clustered needles (Altrogge et al, 2007). Barycenter 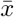 and needle orientation *ā* corresponds to 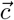 and ***θ***, respectively. Note that the 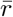 is not the same as *r*.

We note that since covering the boundary target voxels within an ellipse or sphere also covers the internal voxels, we can reduce the constraints in models NOO-MVCE and NOO-MVCS by only considering the boundary target voxels, i.e., *j* ∈𝒯 ′, which we obtain with a grassfire algorithm (Blum, 1967).

#### 2.1.2 Multiple needle placement

For multiple non-parallel *k* needle placement, we first divide tumor voxels into *k* clusters and then identify needle orientation by fitting an ellipse around each cluster using NOO-MVCE (Figure 3). The methodology is referred to as NOO-Kmeans. For a set of *k* needles, we use the classical k-means clustering optimization model to identify these clusters:

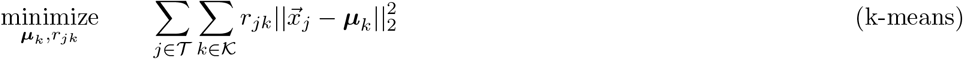

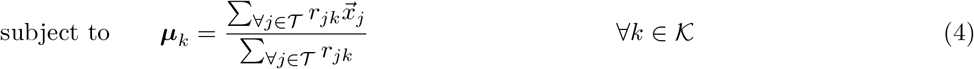

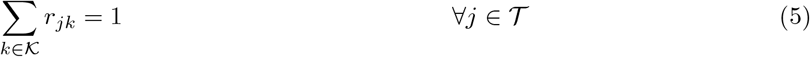

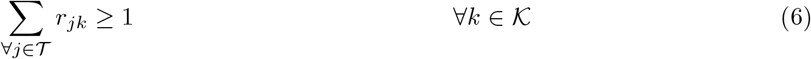

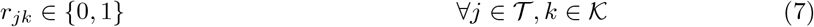

where ***µ***_*k*_ is the mean of cluster *k* and *r*_*jk*_ ∈ {0, 1} indicates if voxel *j* is in cluster *k*. The objective is to minimize the Euclidean distance of voxels from the center (***µ***_*k*_), thereby assigning a voxel to a cluster *k* whose center is closest of all clusters. Constraint 4 computes the mean of a cluster, and Constraint 5 assigns each target voxel to a cluster. Constraint 6 ensures a non-empty cluster. The k-means model is a nonconvex model due to the bilinear term in Constraint 4 as well as in the objective function. A 0-1 SDP-relaxation of k-means has been proposed (Peng and Wei, 2007), but it is intractable for our problem size where the case sizes range from approximately 900 to 62,000 target voxels. Therefore, we solve k-means with Lloyd’s algorithm, which iteratively assigns each voxel to the nearest cluster while updating the centroids until convergence (Lloyd, 1982).

**Figure 3:**
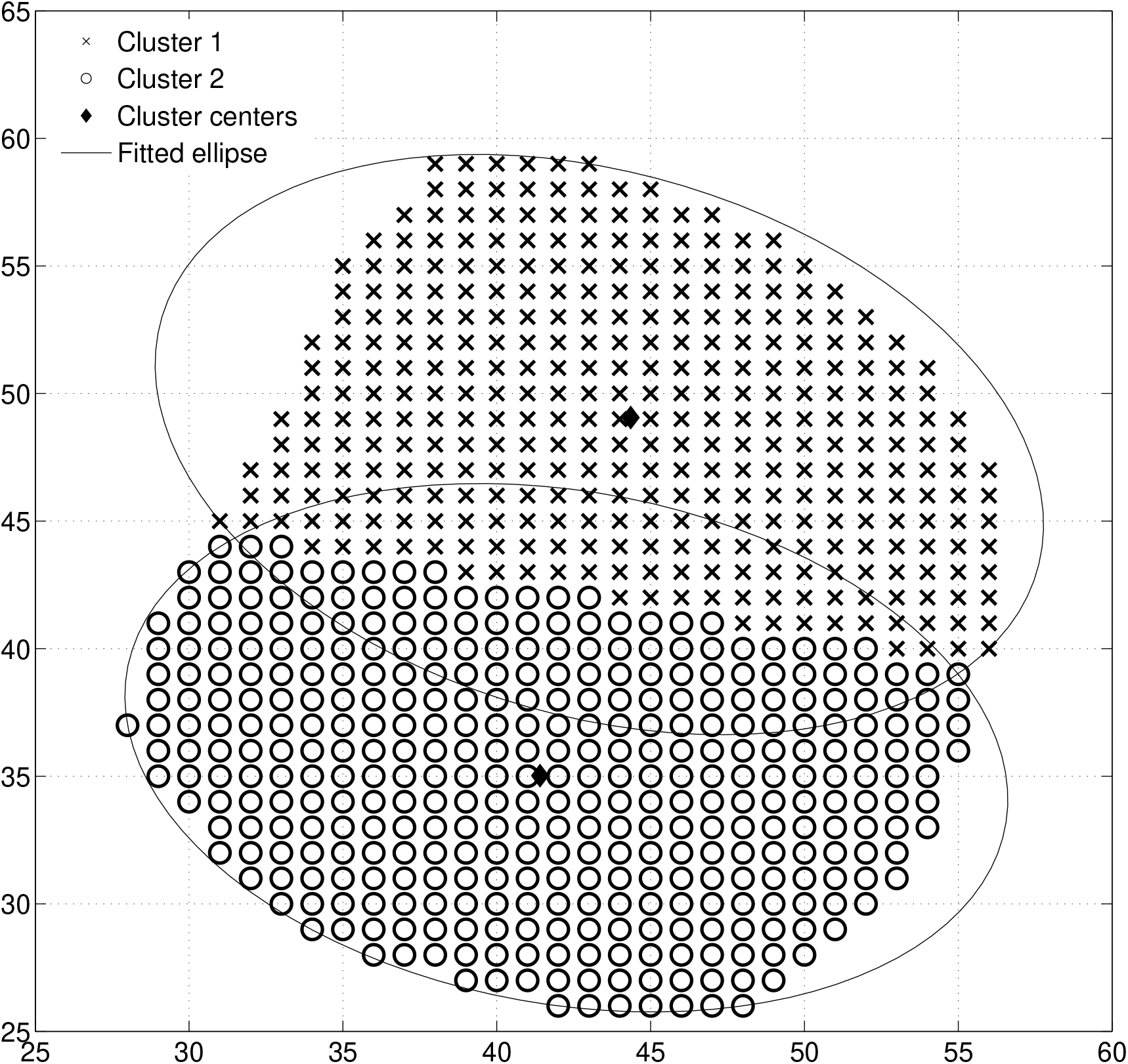
Multiple needle placement using NOO-Kmeans

### 2.2 Thermal dose optimization

Once needle positions are known, we compute the optimal treatment time for adequate thermal dose delivery. We lift the geometric assumption made on the shape of thermal lesion in NOO and compute the actual thermal dose received by the target using BHTE and ATDM thermal damage models. Unlike BHTE which provides temperature at a given time, the ATDM (Henriques, 1947; Henriques and Moritz, 1947; Moritz, 1947; Moritz and Henriques, 1947) thermal damage model considers thermal history of a voxel, i.e., how long a voxel is exposed to a given temperature, and compute cumulative thermal damage over a period of time.

BHTE describes the relationship between tissue local interactions and heat delivery, and is given by the following equation in a 3D system (Ahmed and Goldberg, 2004; Wissler, 1998):

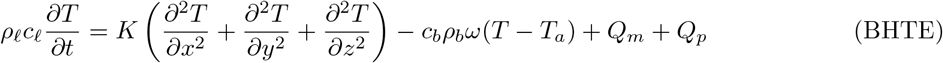

where *ρ* _*𝓁*_ and *ρ*_*b*_ are the densities of tissue and blood (kg/m^3^), respectively; *c*_*𝓁*_ and *c*_*b*_ are the specific heats of the tisue and blood (J/kg-K), respectively; *K* is the thermal conductivity of the tissue (W/m-K); *ω* is the blood perfusion coefficient, i.e., blood flow rate/unit mass tissue (1/s); *T* and *T*_*a*_ are the temperatures of tissue and arterial blood (K), respectively; *Q*_*p*_ is the power absorbed per unit volume of the tissue (W/m^3^); and *Q*_*m*_ is metabolic heating, which is usually considered negligible (Chang and Beard, 2002). The values used for the biological constants and other parameters are given in Table 1. The solution of BHTE gives the temperature of each voxel at each time step.

**Table 1:** Parameter values for BHTE and ATDM

| Parameter | Value |
| --- | --- |
| Blood density ($\rho_b$) (Tungjithkusolmun et al, 2002) | 1000 kg/m<sup>3</sup> |
| Blood heat capacity ($c_b$) (Tungjithkusolmun et al, 2002) | 4180 J/kg-K |
| Blood thermal conductivity (Tungjithkusolmun et al, 2002) | 0.543 W/m-K |
| Liver density ($\rho_\ell$) (Tungjithkusolmun et al, 2002) | 1060 kg/m<sup>3</sup> |
| Liver heat capacity ($\rho_\ell$) (Tungjithkusolmun et al, 2002) | 3600 J/kg-K |
| Liver thermal conductivity ($K$) (Tungjithkusolmun et al, 2002) | 0.512 W/m-K |
| Liver electrical conductivity ($\sigma_\ell$) (Tungjithkusolmun et al, 2002) | 3.33E-3 mS/cm |
| Blood perfusion ($\omega$) (Ebbini et al, 1988) | 6.4E-3 1/s |
| Arterial temperature ($T_a$) | 310.15 K |
| Frequency factor ($A$) (Schwarzmaier et al, 1998) | 3.1E98 1/s |
| Activation energy ($E_A$) (Schwarzmaier et al, 1998) | 6.28E5 J/mo |
| Universal gas constant ($R$) (NIST, 2015) | 8.3145 J/K-mol |

The heat source, *Q*_*p*_, is approximated by (Chen et al, 2006, 2009)

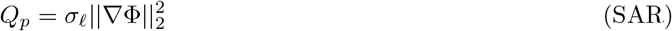

where *σ*_*𝓁*_ is the electrical conductivity of the tissue and Φ is the electric potential. We obtain the electric potential using the Laplacian equation with constant electrical conductivity (Chang and Nguyen, 2004) as follows:

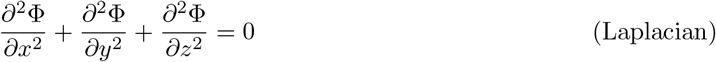

The needle is positioned so that the center of its conducting part is placed at the ellipse or sphere center obtained from NOO. The voxels in contact with the needle are computed from a ray tracing algorithm (Amanatides and Woo, 1987) and form a needle-voxel intersection set. For Laplacian, the initial conditions (voltage) are set to 0 for all voxels except the needle-voxel intersection set, whose initial conditions are set to input voltage of the needle. Both BHTE and Laplacian are solved using a finite difference scheme with homogeneous Neumann boundary conditions (Appendix A).

The Arrhenius thermal damage index is a dimensionless number Ω_*js*_ computed for every voxel *j* of structure *s* and may be interpreted as the probability that the tissue is irreversibly damaged (Pearce, 2009). ATDM is defined as

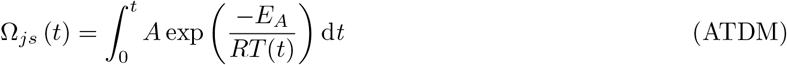

where *A* is the frequency factor, *E*_*A*_ is the activation energy, and *R* is the universal gas constant (Table 1). *T* (*t*) is the average tissue absolute temperature (i.e., temperature in Kelvin) in the time interval [0, *t*] and is obtained from BHTE. Physically, Ω_*js*_ is a natural log of the ratio of the original concentration of undamaged molecules to those at the end of the heating (Pearce, 2009):

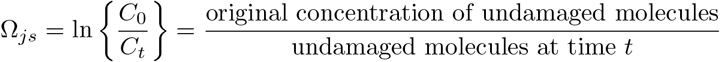

Thus, if *C*_*t*_ ∈ [0, 1] and *C*_0_ = 1, then percentage (or probability) of damaged molecules at time *t* is

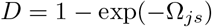

because exp(−Ω_*js*_) = *C*_*t*_ = undamaged molecules at time *t*. We describe these percentage damage models as *D*63 for *p* = 63% tissue damage, *D*70 for *p* = 70% tissue damage, etc. A value of *p* = 0.63 or 63% is associated with irreversible thermal damage and corresponds to Ω_*js*_ = 1.

We define a set of needle configurations as a combination of needle type (*n* ∈𝒩) and source voltage (*ϕ*∈𝒱). The set of damage models is given by: *d* ∈𝒟 = {BHTE, ATDM, D63, D70, D80, D95}. For each needle configuration, (*ϕ, n*), we first compute the BHTE for fixed treatment time using inputs from Laplacian and then compute the ATDM followed by the percentage damage models if required for thermal damage *d*. We save this information to determine the minimum treatment voltage and treatment times. We define a numerical dose structure 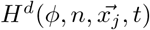 to identify damage using model *d* to voxel 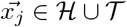 at time *t* due to needle configuration (*ϕ, n*).

For a fixed treatment time *t*_max_, the minimum treatment voltage for full target coverage using damage model *d* ∈𝒟 and needle type *n* ∈𝒩 is given by

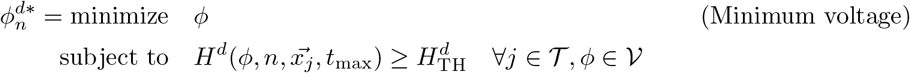

where 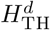 is the threshold damage value for model *d*, 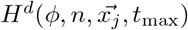 is the damage to voxel 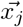 at *t*_max_ minutes when using needle configuration (*ϕ, n*). If temperature is used to quantify thermal damage, then 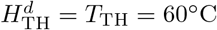; if the Arrhenius damage index is used to quantify thermal damage, then 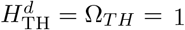; and if *p* percent damage is used to quantify thermal damage, then 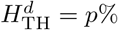.

Similarly, for a fixed voltage *ϕ*, the minimum treatment time for full target coverage using damage model *d* ∈𝒟 and needle type *n* ∈𝒩 is given by

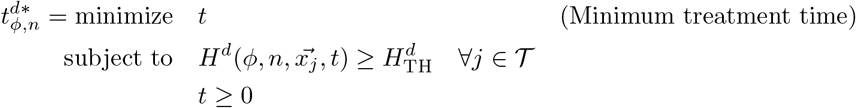

To choose a single best needle configuration for damage model *d*, we select the needle configuration with 100% coverage and the least OAR damage (Algorithm 2).

## 3 Results

We perform all computations on Intel Core i7-3770 CPU with 3.40 GHz and 8 GB RAM using MATLAB R2015b (Mathworks, Inc.). Our case studies are liver cases (Table 2) obtained from Robarts Research Institute, Western University. In a clinical setting, tumors are over-ablated to ensure microscopic tumor particles are killed along with the target itself. Therefore, we add surgical margins of 0 mm (N), 3 mm (S), 5 mm (M), 10 mm (L) around the target (Table 2). Further, in liver ablation OAR sparing is insignificant due to its regenerative properties and hence no explicit OAR margin is added to the target. However, we consider damage to non-target voxels outside surgical margin as OAR damage to understand the impact of input parameters.

**Table 2:** Description of case studies

| ID | Volume<br>(mm<sup>3</sup>) |  | Target<br>voxels |  | Boundary<br>target voxels |  |
| --- | --- | --- | --- | --- | --- | --- |
|  | 100% | 50% | 100% | 50% | 100% | 50% |
| 1N | 898 | 544 | 898 | 544 | 501 | 293 |
| 1S | 3063 | 1641 | 3063 | 1641 | 840 | 430 |
| 1M | 5138 | 2595 | 5138 | 2595 | 1106 | 808 |
| 1L | 13003 | 5874 | 13003 | 5874 | 1911 | 1839 |
| 2N | 4657 | 2090 | 4657 | 2090 | 1591 | 738 |
| 2S | 10595 | 4430 | 10595 | 4430 | 2178 | 747 |
| 2M | 15595 | 6320 | 15595 | 6320 | 2610 | 1058 |
| 2L | 32225 | 12327 | 32225 | 12327 | 3830 | 2084 |
| 3N | 13481 | 5183 | 13481 | 5183 | 3273 | 1334 |
| 3S | 24895 | 9169 | 24895 | 9169 | 4060 | 1816 |
| 3M | 33881 | 12226 | 33881 | 12226 | 4640 | 2158 |
| 3L | 61771 | 21442 | 61772 | 21442 | 6230 | 3070 |

### Algorithm 2

Best needle configuration

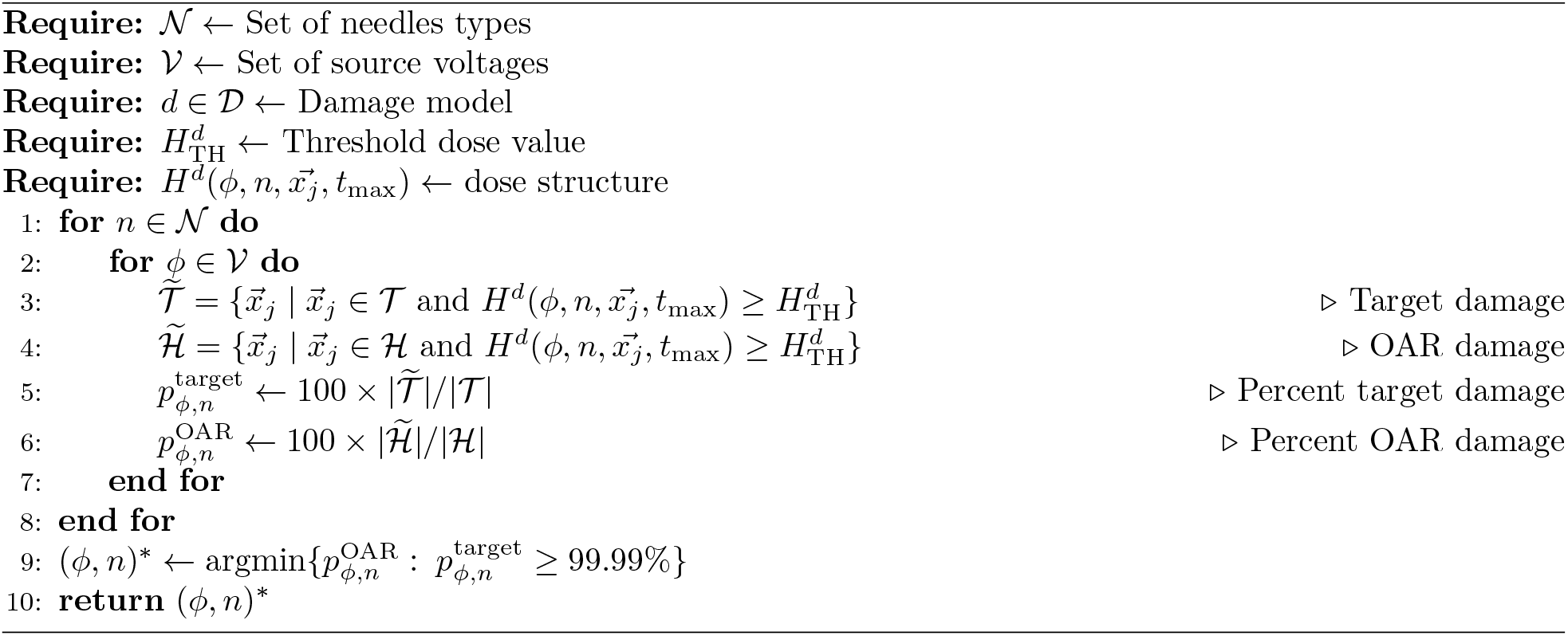

### 3.1 NOO results

We use CVX (CVX Research, 2012; Grant and Boyd, 2008) to solve NOO-MVCE and NOO-MVCS models and test our NOO approach under two scenarios, (1) considering all the target voxels, and (2) considering only boundary target voxels, obtained using a grassfire algorithm (Blum, 1967). For faster computation, both these scenarios are solved for the entire target as well as targets sampled at 50%. We restrict our needle types, 𝒩, to eight Covidien specifications (Table 3) (Medtronic, 2016).

**Table 3:** Needle types (𝒩)

| Abbreviation | Needle description | Active tip length (mm) |
| --- | --- | --- |
| SN7 | Single needle | 7 |
| SN10 | Single needle | 10 |
| SN20 | Single needle | 20 |
| SN30 | Single needle | 30 |
| CN25 | Clustered needle | 25 |
| MN2K30 | Multiple needle, $k = 2$ | 30 |
| MN3K30 | Multiple needle, $k = 3$ | 30 |
| MN3K40 | Multiple needle, $k = 3$ | 40 |

As the number of voxels increases, the runtimes for both NOO-MVCE and NOO-MVCS increase (Figure 4). The runtimes of NOO-MVCS are under 8 seconds for all unsampled target voxels and under 3 seconds for unsampled boundary target voxels (Figure 4(a)). Using boundary target voxels gives an average computational gain of 60% and 53% for NOO-MVCS for unsampled and sampled case, respectively (Figures 4(a) and 4(b)). For all unsampled target voxels, NOO-MVCE does not finish in reasonable amount of time (*>*1 hour) for Cases 2L and 3M, while Case 3L runs out of memory (Figure 4(c)). However, NOO-MVCE runs in under a minute for unsampled boundary target voxels in all cases. An average computational gain of 81% and 83% is obtained for unsampled and sampled cases, respectively, when only boundary voxels are used for NOO-MVCE (Figures 4(c) and 4(d)).

**Figure 4:**
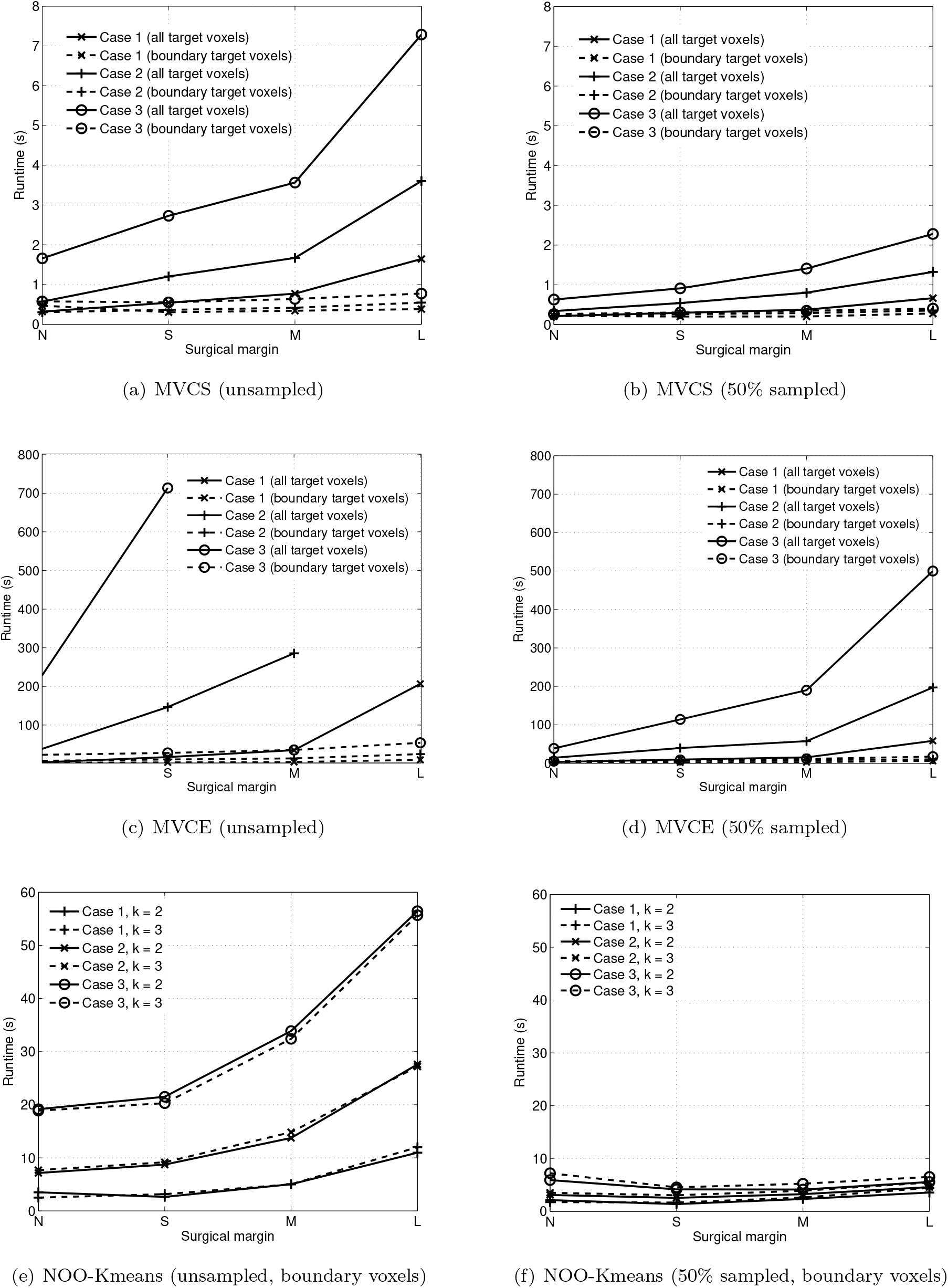
NOO Runtimes

We use boundary voxels to solve MVCE for NOO-Kmeans since using boundary voxels only has significant computational advantage demonstrated earlier (Figures 4(e) and 4(f)). For unsampled cases, NOO-Kmeans runs under a minute (Figure 4(e)). These fast runtimes may appear counter-intuitive since NOO-MVCE is solved *k* times, once for each cluster. However, each cluster contains only a subset of target voxels, and we consider only the boundary voxels of these clusters.

When using boundary voxels, runtimes are under a minute for the largest unsampled case (Case 3L). Therefore, we report results only for unsampled cases for both NOO and TDO. For selected cases, Figure 5 shows the needle orientations given by MVCE, MVCS, and NOO-Kmeans models, and Table 4 shows their fitted volumes.

**Figure 5:**
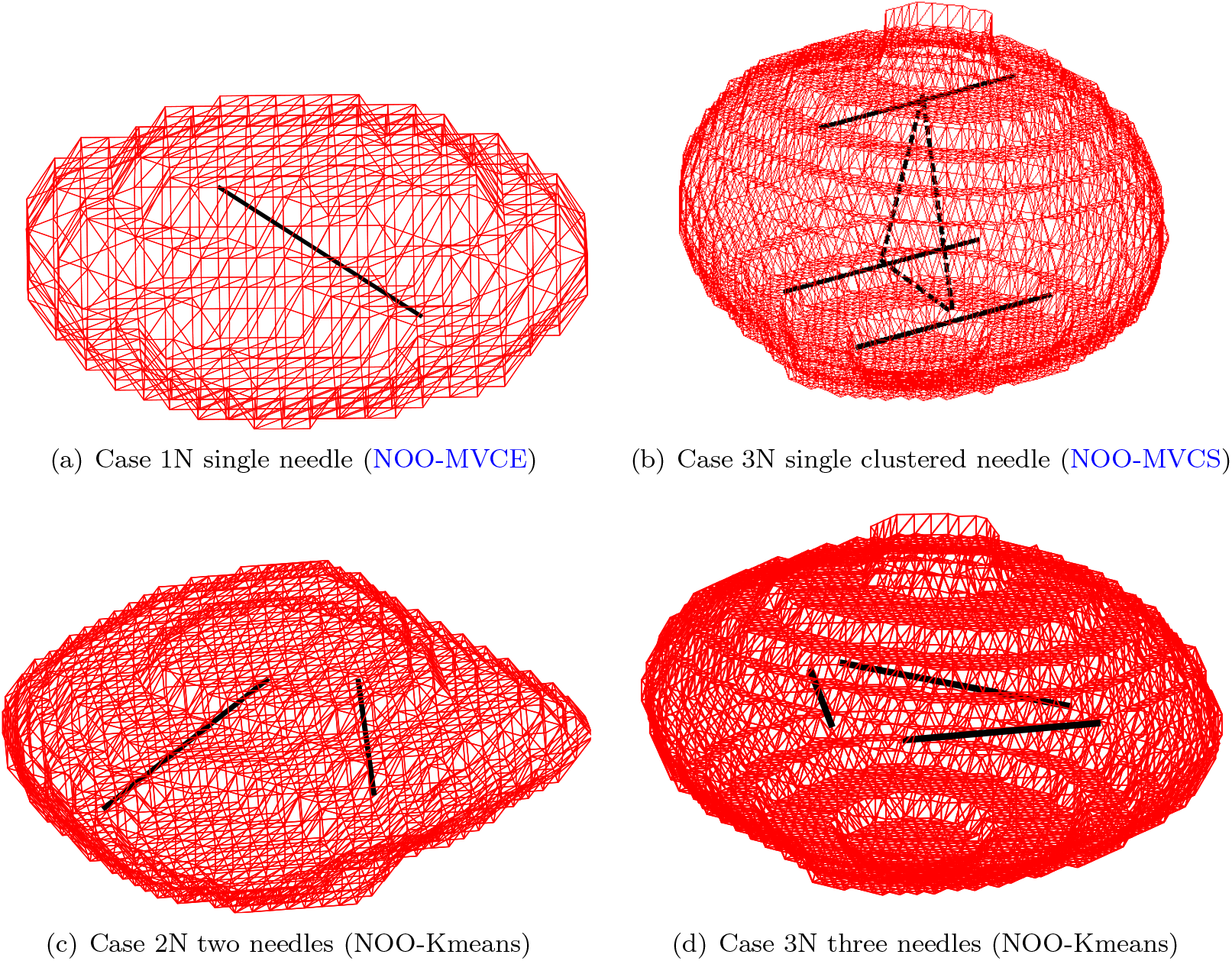
Needle positions and orientations. The dotted lines in 5(b) shows the equilateral triangle whose vertices correspond to the centers of the conducting tines in the cluster.

**Table 4:** Numerical results for NOO 𝒜 (*ξ*) = Fitted volumes, 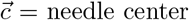, ***θ*** = needle orientation

| ID | $\mathcal{A}(\xi)$ (mm<sup>3</sup>) | $\vec{c}$ | $\boldsymbol{\theta}$ | Model |
| --- | --- | --- | --- | --- |
| 1N | 956 | $[35.24 \ 34.39 \ 47.15]^\top$ | $[-0.87 \ -0.5 \ 0.03]^\top$ | NOO-MVCE |
| 2N | $\begin{bmatrix} 3922.6 \\ 3929.6 \end{bmatrix}$ | $\begin{bmatrix} 41.23 & 36.67 & 40.68 \\ 43.35 & 46.95 & 40.19 \end{bmatrix}^\top$ | $\begin{bmatrix} -0.1 & 0.1 & 0.03 \\ 0.82 & -0.57 & -0.01 \end{bmatrix}^\top$ | NOO-Kmeans |
| 3N | 48419 | $[47.79 \ 48.17 \ 69.59]^\top$ | $[1 \ 0 \ 0]^\top$ | NOO-MVCS |
| 3N | $\begin{bmatrix} 9881.4 \\ 9536.4 \\ 9519.6 \end{bmatrix}$ | $\begin{bmatrix} 42.62 & 54.58 & 70.39 \\ 55.87 & 50.00 & 70.14 \\ 45.65 & 39.90 & 70.45 \end{bmatrix}^\top$ | $\begin{bmatrix} 0.8 & 0.59 & -0.06 \\ 0.16 & -0.99 & 0.03 \\ -0.94 & 0.39 & 0.02 \end{bmatrix}^\top$ | NOO-Kmeans |

**Table 5:** Case 1N results for *n* = SN7

| Damage model ($d$) | $\phi_n^{d*}$ | $t_{\phi,n}^{d*}$ (min) |
| --- | --- | --- |
| BHTE | 20.00 | 1.20 |
| ATDM | 25.00 | 1.94 |
| D63 | 25.00 | 1.93 |
| D70 | 25.00 | 2.20 |
| D80 | 27.50 | 1.90 |
| D95 | 27.50 | 2.27 |

### 3.2 TDO results

We solve BHTE and Laplacian using a finite difference scheme (Appendix A) to obtain thermal distributions over a 20 min simulation with a 0.5 times step for eight needle types (Table 3) with source voltages, 𝒱, varying from 2.5 V to 30 V in increments of 2.5 V. Thermal distributions are computed only if non-intersecting needle positions are found. Each case consists of 384 runs: 8 needle types ×4 surgical margins ×12 source voltages. The computational runtime of each run is the total time to solve Laplacian, BHTE, and ATDM; computational time is largely driven by the Laplacian (Figure 6(a)). We assess target and OAR damage using the following thermal damage models: (1) ≥60^°^C threshold temperature from BHTE (T60), (2) Arrhenius damage model (ATDM), (3) 63% damage (D63), (4) 70% damage (D70), (5) 80% damage (D80), and (6) 95% damage (D95).

**Figure 6:**
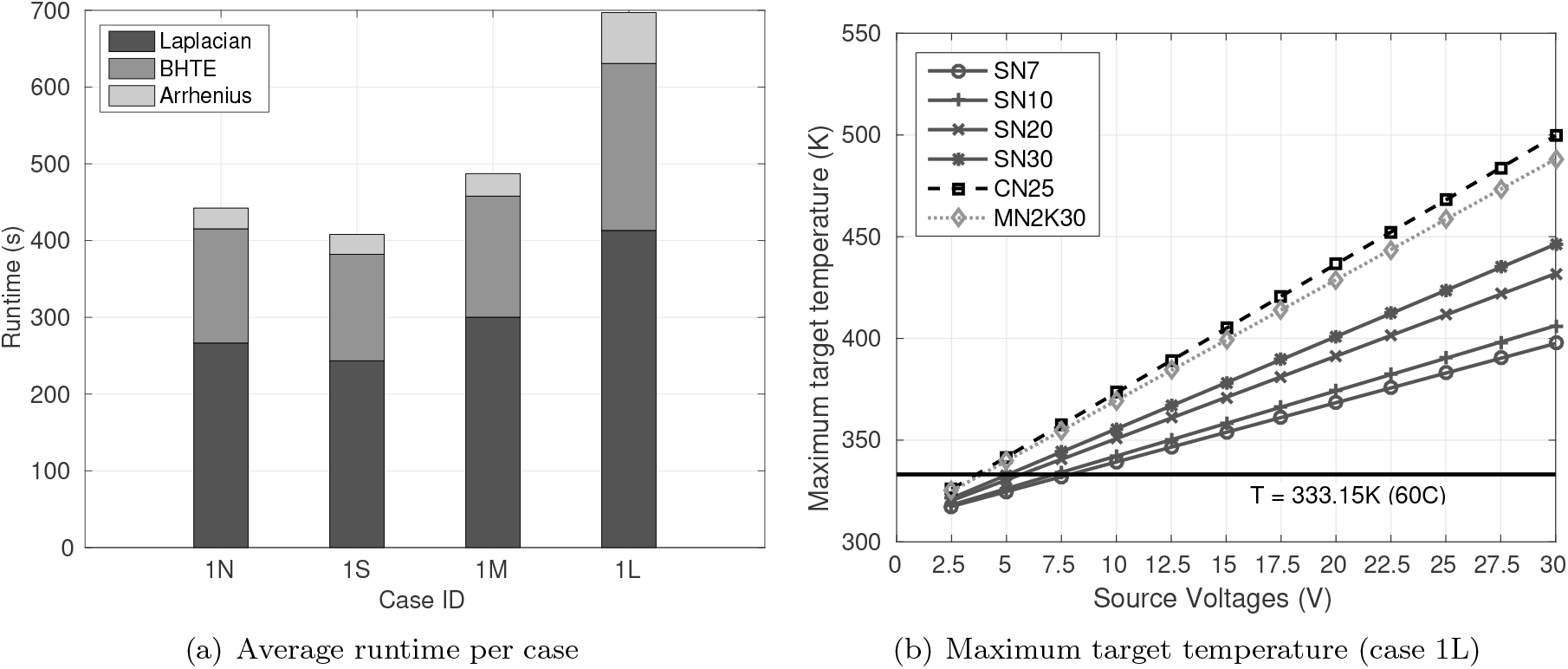
Average runtimes and maximum target temperature

The maximum temperature in the target increases with an increase in source voltage; at least 7.5V is recommended for the representative target treatment (Figure 6(b)). High source voltage increases the numerical value of the initial conditions for the Laplacian, causing high target temperatures, while longer or multiple needles increases the needle-voxel intersection set, resulting in larger thermal spread. Hence, more needles or high source voltage yields large ablation volumes (Figure 7) and high target (Figure 8) and OAR damage (Figure 9), and consequently high tissue molecular damage.

**Figure 7:**
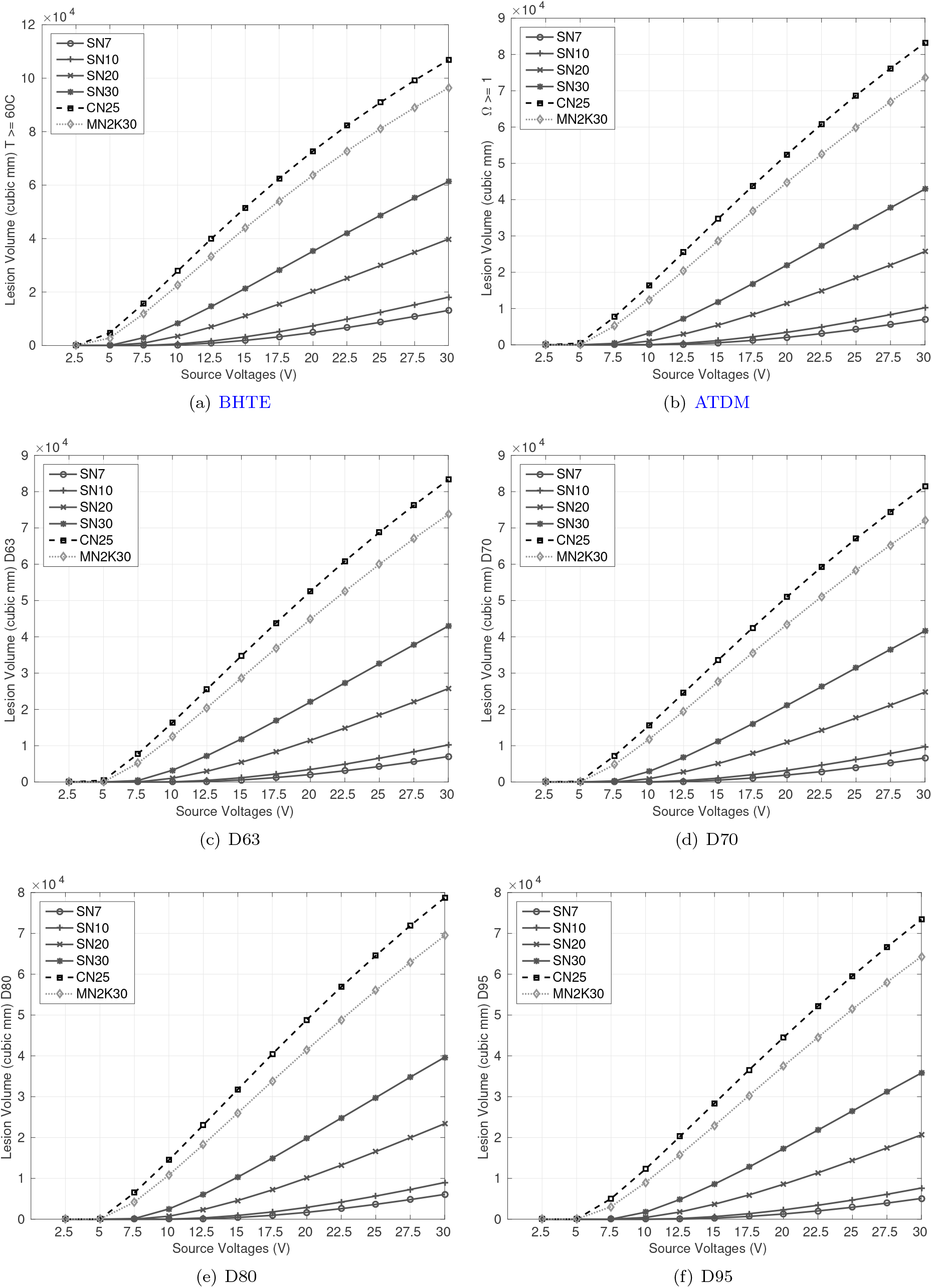
Lesion volumes (Case 1N)

**Figure 8:**
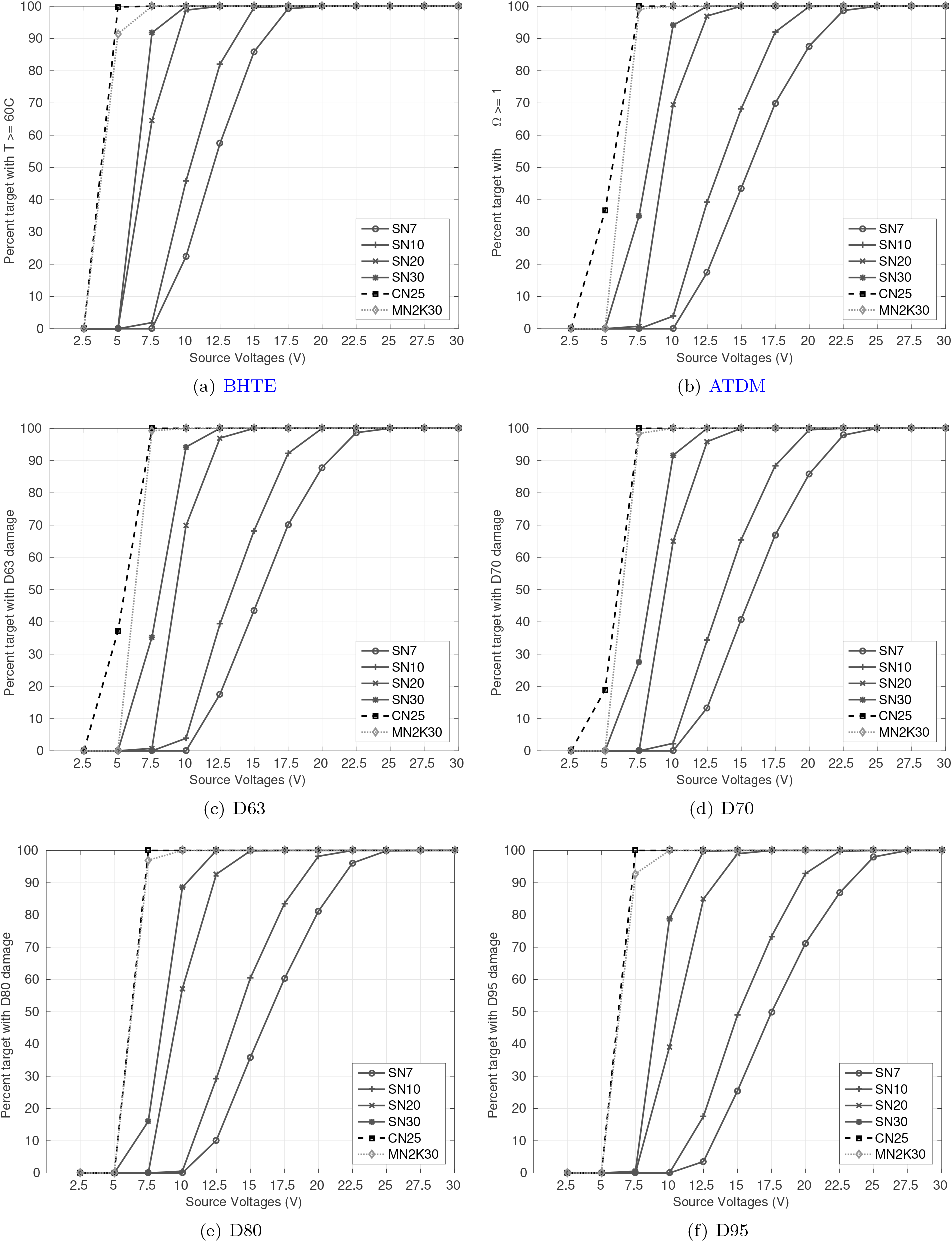
Percent target coverage (Case 1N)

**Figure 9:**
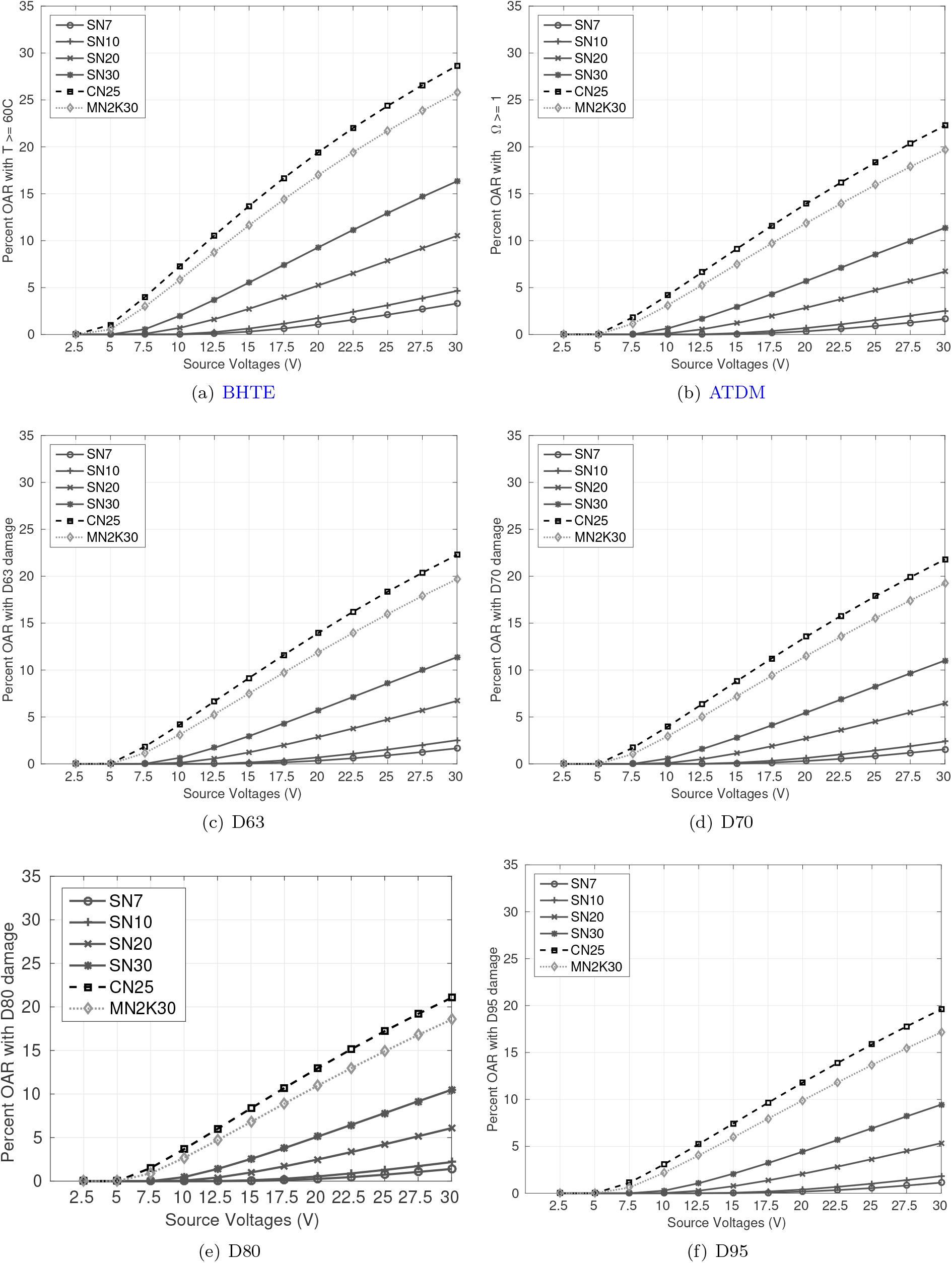
Percent OAR coverage (Case 1N)

Full coverage is seen when more needles operate at low voltage or fewer needles operate at high voltage. Further, a low and high source voltage is recommended when damage is quantified by BHTE and D95 models, respectively, resulting in a different needle configurations for the same case. This difference in needle configuration arises because tissue molecular damage increases with the duration of exposure to temperatures ≥60^°^C, and BHTE damage occurs before D95 damage (Figure 10). Therefore, certain needle configurations achieve full BHTE coverage but partial D95 coverage because all the target voxels are not exposed long enough at temperatures ≥60^°^C. Thus, BHTE damage requires a low source voltage and high tissue molecular damage requires high source voltage (Table 6). Finally, our framework indicates the use of a single needle for targets up to 15 cm^3^ and multiple needles for larger targets (Figure 11). Needle configurations that do not attain full coverage are not recommended for treatment.

**Figure 10:**
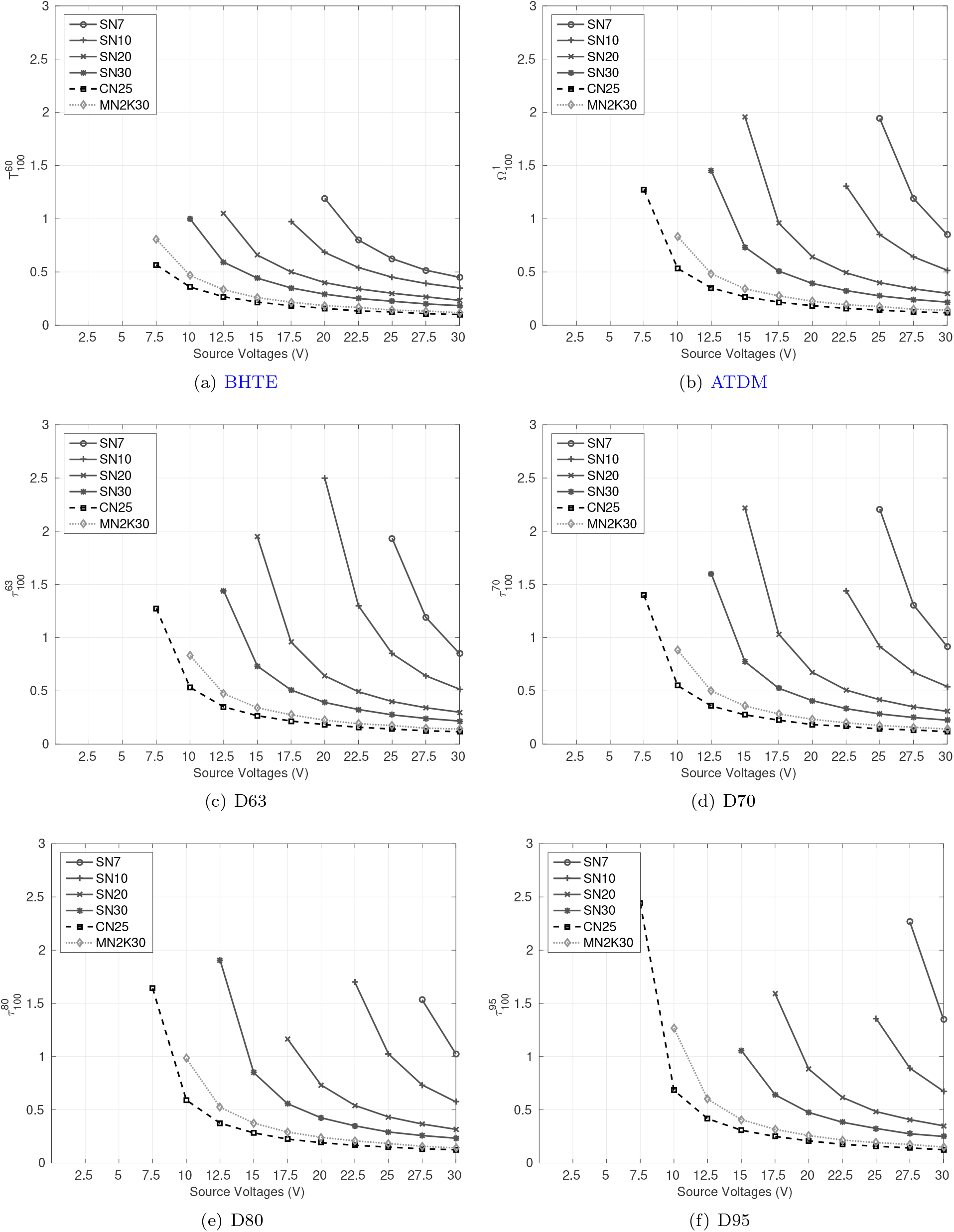
Treatment times, rounded to the closest minute, based on full coverage (Case 1N)

**Table 6:** Recommended needle configurations (Algorithm 2)

| Case ID | Damage model | Needle type | Volts (V) | OAR damage (%) |
| --- | --- | --- | --- | --- |
| 1N | BHTE | SN7 | 20.00 | 1.07 |
| 1N | ATDM | SN7 | 25.00 | 0.91 |
| 1N | D63 | SN10 | 20.00 | 0.69 |
| 1N | D70 | SN7 | 25.00 | 0.84 |
| 1N | D80 | SN10 | 22.50 | 0.89 |
| 1N | D95 | SN7 | 27.50 | 0.82 |
| 2N | BHTE | MN2K30 | 7.50 | 2.21 |
| 2N | ATDM | MN2K30 | 10.00 | 2.34 |
| 2N | D63 | MN2K30 | 10.00 | 2.35 |
| 2N | D70 | MN2K30 | 10.00 | 2.16 |
| 2N | D80 | SN20 | 25.00 | 2.42 |
| 2N | D95 | SN20 | 27.50 | 2.62 |
| 3N | BHTE | MN2K30 | 27.50 | 6.03 |
| 3N | ATDM | MN3K30 | 30.00 | 8.85 |
| 3N | D63 | MN3K30 | 30.00 | 8.85 |
| 3N | D70 | MN3K30 | 30.00 | 8.57 |
| 3N | D80 | MN3K30 | 30.00 | 8.15 |
| 3N | D95 | - | - | - |

**Figure 11:**
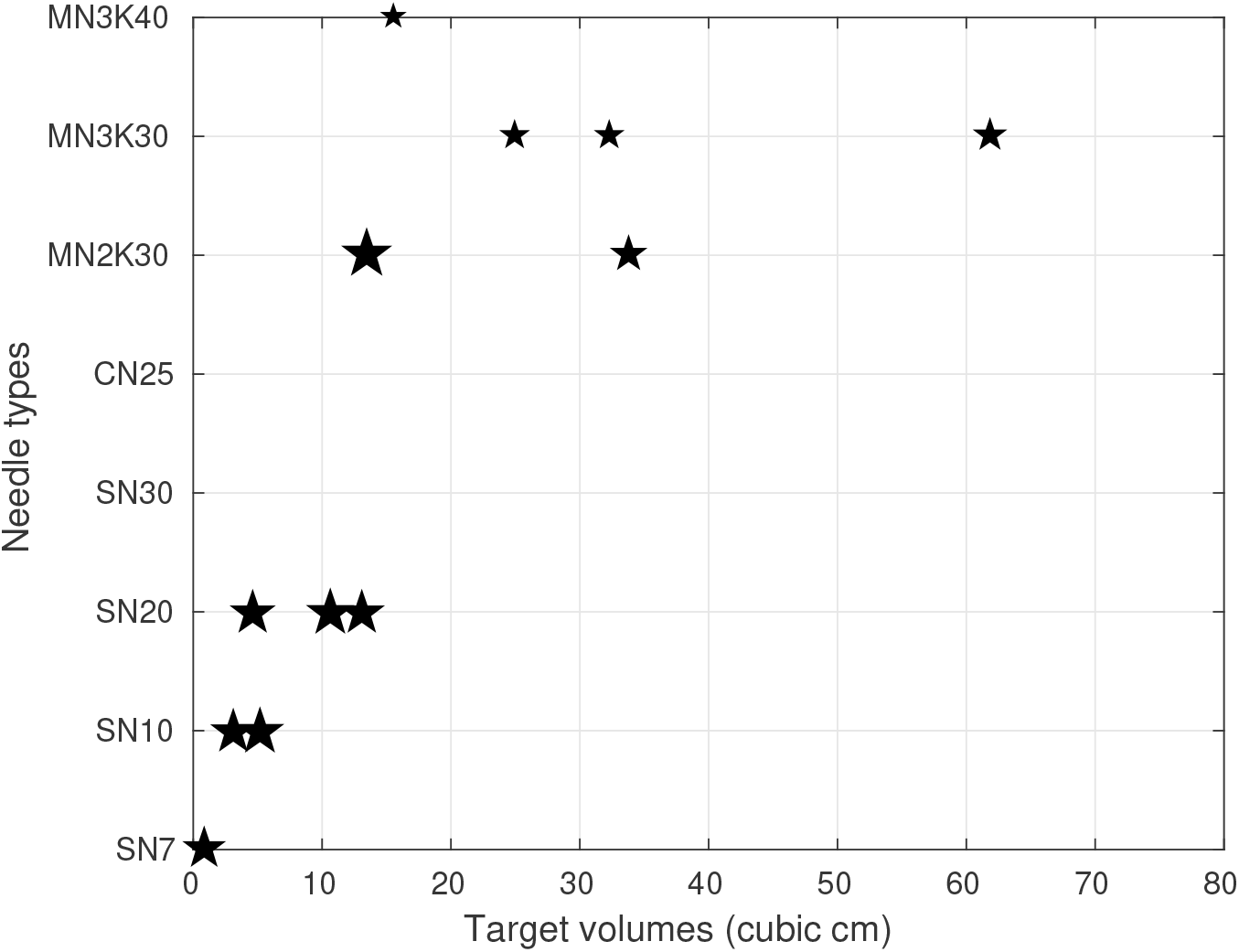
Recommended needle configuration for BHTE damage model. The voltage is indicated by the star size.

Our multiple needle placement methodology is unable to find non-intersecting needle positions for smaller tumors using longer multiple needles (e.g., MN3K30, MN3K40 for Case 1N), and hence no TDO computations were performed for such cases. However, for Case 3N, NOO-Kmeans could not find non-interesting needle positions for MN3K40, and none of the other needle configurations were able to obtain 100% target coverage. In such circumstance we increase the target size by adding margins to obtain needle positions and perform TDO analysis for the original target.

## 4 Discussion

Due to the lack of standards based on either conformity or OAR-sparing, complexity of optimization models, and difference in data sets and needle types used, it is difficult to draw direct comparisons with existing simultaneous models. In ablation planning, simultaneous optimization provides the benefit of needle placement by simultaneously computing thermal damage without any assumptions on ablation shape. Due to inherent non-linear nature of ablation, simultaneous optimization methods, that solve PDEs as constraints with needle position and orientation as only variables, are only able to produce locally optimal solutions. They must be tailored to needle type as well as ablation modality thereby restricting their clinical viability. Trajectory planning is difficult to incorporate in such models and due to long run-times experiments on multiple source voltage (or power) selection is not tractable. Further, due to the mathematical complexity of ablation optimization models, it is difficult to comment on the quality of their optimal solutions and none of the existing models comment on optimality gaps. Decomposing needle placement and thermal damage computation, as proposed in this work, results in inherent sub-optimal solutions. However, such an approach provides several benefits, including computational advantage, flexibility towards ablation modality, flexibility to incorporate trajectory planning, and needle types, that is not seen in simultaneous models.

Typically, a good cancer treatment plan will provide a full conformal target coverage with maximum OAR sparing. However, unlike radiation, rigidity of heat deposition makes it difficult to control the shape or spread of ablation. If full target coverage is the only necessary requirement, then any needle position that achieves this goal is an acceptable solution. However, it is obvious that some needle positions are better than others. For instance, a needle, that is larger than target radius, placed closer to target boundary may provide full coverage but is less desirable than one closer to the center of the target. This choice can be attributed to better target thermal dose, coverage of microscopic tumor particles surrounding the target, and less OAR damage. Existing models do not provide information on OAR sparing and use different data sets for any comparative analysis and we did not find any standard in the literature to evaluate quality of an RFA treatment based on either OAR-sparing or target conformity.

Intuitively, for a single needle placement, the needle position will correspond to the centroid of the target and its orientation will correspond to the shape of the target. This hypothesis has been previously validated through experiments using simultaneous optimization (Altrogge et al, 2007). Our fast convex NOO model, NOO-MVCE, is able to deliver similar solutions. The grassfire method to extract boundary voxels yields NOO solutions in *<*1 minute. For multiple needle placement, we provide detailed methodological explanation absent in previous work using same approach (Chen et al, 2009).

Any geometric assumptions in NOO stage are lifted in TDO stage and actual thermal dosimetry is computed using several damage models. The thermal dose for the largest target (Case 3L) is computed in *<*20 minutes, which is a significant improvement over the 1-2 hours reported by simultaneous optimization (Altrogge et al, 2007; Chen et al, 2009). We can easily extend our work to other ablation modalities by solving a different set of PDEs, e.g., Maxwell’s equations for MWA, which can be difficult in a PDE-constrained systems. Further, new needle types can be seamlessly added in the NOO stage without affecting the methodology for thermal dose computations.

Similar to simultaneous optimization methods, we assume a fixed treatment time which in our case is 20 minutes. This conservative longer treatment time gives us enough simulation data to analyze the treatment quality while ensuring maximum target coverage. However, for the recommended needle configurations, full coverage is achieved within the first few minutes. Since tissues eventually reach thermal equilibrium, treatment time does not significantly affect the treatment quality, unlike radiation treatments. Gradual heat deposition ensures larger ablation volumes and therefore the coverage of microscopic tumor particles with longer treatment time increasing the tissue molecular damage.

We addressed models for design ablation treatments in a deterministic setting, where needle placements are exact. However, ultrasound image guidance may be inaccurate up to 2.5 mm (Neshat et al, 2013), and further, needles may deflect (bend) unexpectedly during insertion. Clinicians can often counteract deflection when it is observed, however, the frequency and causes of deflection are not well understood, though there are efforts to anticipate and estimate needle deflection (e.g., Jiang et al (2018)). We note that despite these uncertainties, clinical treatments, like our models presented here, are not designed with these uncertainties in consideration. We also note that our decoupled NOO and TDO approach sacrifices an unknown amount of objective function quality compared to simultaneous NOO and TDO, but simultaneous optimization of both yields a challenging and intractable model.

## 5 Conclusion and future work

While the current state-of-the-art in ablation planning performs simultaneous optimization, these methods lack in computational tractability and flexibility to accommodate different needle types and ablation modality. In this work, we present a novel systematic approach to radiofrequency ablation where we present a dissociated needle placement and thermal dose computation methodology that can be extended to other ablation modalities (e.g., microwave ablation). We eliminate the need to iteratively compute thermal dose thereby improving the computational tractability of developing a treatment plan. Further, our framework considers NOO and TDO for eight different needle types and is able to accommodate other needles (e.g., umbrella-shaped needles).

We designed fast convex NOO models for single and clustered needles that can be solved to optimality as well as fast heuristic approach for multiple random needle placement. We are also able to compute the Laplacian, BHTE, ATDM, and percent damage models in reasonable amount on time. However, the use of commercial PDE solvers that include RF modules ((COMSOL Inc., 2017)) can enhance the quality of these solutions, and therefore treatment plans, since several physiological, thermal, and electrical process (e.g, change in tissue thermal properties with temperature change) are difficult to capture by mathematical simulations presented here. Further extensions to this work include addressing uncertainties in needle placement and deflection. Although trajectory planning and thermal dose validations must be performed before clinical applicability, we present a promising framework that gives 100% target coverage while, for the first time, performing OAR damage analysis using several different thermal damage models. We analyse the effect of different needle types (needle tip length and number of needles) on target and OAR damage using several damage models when operated under different source voltages. Finally, our methods culminate to the best needle configuration based on full target and minimum OAR coverage.

## Data Availability

Data not available due to patient privacy

## 6 Acknowledgements

The authors would like to thank Hamid Neshat, Western University, for providing data and domain knowledge.

## A Finite difference schemes: Laplacian and BHTE

The electrostatic equation given by Laplacian is solved using an explicit central finite difference scheme as follows:

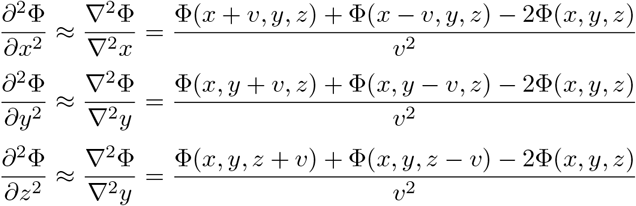

where Φ(*x, y, z*) is the voltage at position (*x, y, z*) and *v* is the dimension of the voxel.

Let Ψ represent the problem domain, and Ψ_*b*_ ⊂ Ψ represent the voxels at the boundary of the domain. 𝒯 ⊂ Ψ be the set of voxels representing the tumor, 𝒯 _*N*_ ⊂𝒯 be the voxels in contact with the needle, and *S* ⊂ Ψ be the voxels representing the OARs.

The initial conditions are given by:

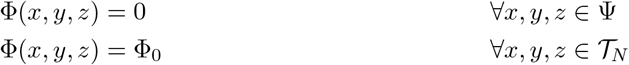

and the homogeneous Neumann boundary conditions Strauss (1992) are given by:

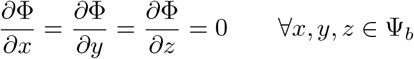

where Φ_0_ is the input voltage.

Pennes’ bioheat equation in 3D given by Equation BHTE is solved using an explicit central finite difference scheme. The temperature is evaluated as follows:

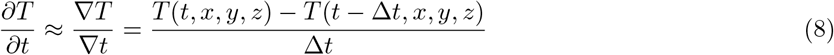

where *T* (*t, x, y, z*) is the temperature of a voxel at position (*x, y, z*) at time *t*, and Δ*t* is the time step length or the frequency in seconds when temperature measures are made. The spatial coordinates are approximated as follows:

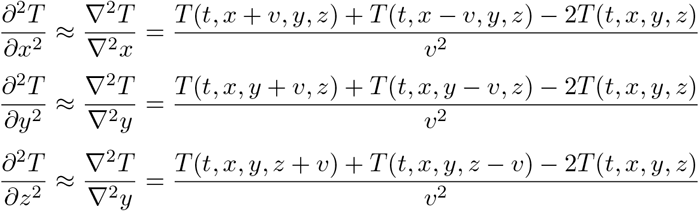

The initial condition is given by:

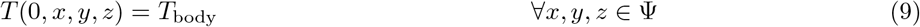

and the homogeneous Neumann boundary conditions Strauss (1992) are given by:

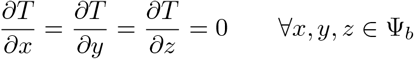

where *τ* is total simulation time, *T*_body_ = 310.15K is the body temperature. We use voxel size 1mm × 1mm × 1mm or *v* = 1mm and the time step is Δ*t* = 0.5s.

## References

Ahmed M, Goldberg SN (2004) Radiofrequency tissue ablation: principles and techniques. In: Radiofrequency Ablation for Cancer, Springer, pp 3–28

Altrogge I, Preusser T, Kroger T, Buskens C, Pereira PL, Schmidt D, Peitgen HO (2007) Multiscale optimization of the probe placement for radiofrequency ablation. Academic radiology 14(11):1310–24, DOI 10.1016/j.acra.2007.07.016, URL http://linkinghub.elsevier.com/retrieve/pii/S1076633207004448

Amanatides J, Woo A (1987) A fast voxel traversal algorithm for ray tracing. In: Eurographics, pp 3–10

Bhardwaj N, Strickland A, Ahmad F, Dennison A, Lloyd D (2010) Liver ablation techniques: A review. Surgical Endoscopy 24:254–265, DOI 10.1007/s00464-009-0590-4, URL http://dx.doi.org/10.1007/s00464-009-0590-4

Blum H (1967) A transformation for extracting new descriptors of shape. Models for the perception of speech and visual form 19(5):362–380

Boyd S, Vandenberghe L (1996) Semidefinite programming. SIAM Review 38(1):49–95

Boyd S, Vandenberghe L (2004) Convex Optimization. Cambridge University Press

Butz T, Warfield S, Tuncali K, Silverman S, Sonnenberg Ev, Jolesz F, Kikinis R (2000) Pre- and intra- operative planning and simulation of percutaneous tumor ablation. International Conference on Medical Image Computing and Computer Assisted Intervention 3:317–326

Chang I, Beard BB (2002) Precision test apparatus for evaluating the heating pattern of radiofrequency ablation devices. Medical Engineering & Physics 24:633–640

Chang I, Nguyen U (2004) Thermal modeling of lesion growth with radiofrequency ablation devices. BioMedical Engineering OnLine 3(1):27, DOI 10.1186/1475-925X-3-27, URL http://www.biomedical-engineering-online.com/content/3/1/27

Chen CCR, Miga M, Galloway R (2006) Optimizing needle placement in treatment planning of radiofrequency ablation. In: Medical Imaging 2006: Visualization, Image-Guided Procedures, and Display, vol 6141, pp 632–638, DOI 10.1117/12.654983, URL http://link.aip.org/link/PSISDG/v6141/i1/p614124/s1&Agg=doi

Chen CCR, Miga MI, Galloway RL (2009) Optimizing electrode placement using finite-element models in radiofrequency ablation treatment planning. IEEE Transactions on Biomedical Engineering 56(2):237–45, DOI 10.1109/TBME.2008.2010383, URL http://ieeexplore.ieee.org/lpdocs/epic03/wrapper.htm?arnumber=4694126

COMSOL Inc (2017) Microwave and RF design software. http://www.comsol.com/rf-module, [Online; accessed 5-January-2017]

CVX Research (2012) CVX: Matlab software for disciplined convex programming, version 2.0 beta. http://cvxr.com/cvx

Dupuy D, Zagoria R, Akerley W, Mayo-Smith W, Kavanagh P, Safran H (2000) Percutaneous radiofrequency ablation of malignancies in the lung. American Journal of Roentgenology 174(1):57–59

Ebbini E, Umemura SI, Ibbini M, Cain CA (1988) A cylindrical-section ultrasound phased-array applicator for hyperthermia cancer therapy. IEEE Transactions on Ultrasonics, Ferroelectrics and Frequency Control 35(5):561–572, DOI 10.1109/58.8034

Eggener S, Scardino P, Carroll P, Zelefsky M, Sartor O, Hricak H, Wheeler T, Fine S, Trachtenberg J, Rubin M, Ohori M, Kuroiwa K, Rossignol M, Abenhaim L (2007) Focal therapy for localized prostate cancer: A critical appraisal of rationale and modalities. The Journal of Urology 178(6):2260 – 2267, DOI http://dx.doi.org/10.1016/j.juro.2007.08.072, URL http://www.sciencedirect.com/science/article/pii/S0022534707021623

Ferris M, Shepard DM (2000) Optimization of gamma knife radiosurgery. Discrete Mathematical Problems with Medical Applications 55:27–44

Ferris M, Lim J, Shepard D (2002) An optimization approach for radiosurgery treatment planning. SIAM Journal on Optimization 13(3):921–937, DOI 10.1137/S105262340139745X, URL http://dx.doi.org/10.1137/S105262340139745X, http://dx.doi.org/10.1137/S105262340139745X

Ghobadi K, Ghaffari H, Aleman D, Jaffray D, Ruschin M (2012) Automated treatment planning for a dedicated multi-source intracranial radiosurgery treatment unit using projected gradient and grassfire algorithms. Medical Physics 39(6):3134–3141, DOI 10.1118/1.4709603

Ghobadi K, Ghaffari H, Aleman D, Jaffray D, Ruschin M (2013) Automated treatment planning for a dedicated multi-source intra-cranial radiosurgery treatment unit accounting for overlapping structures and dose homogeneity. Medical Physics 40(9)

Goldberg SN, Gazelle S, Mueller P (2000) Thermal ablation therapy for focal malignancy: a unified approach to underlying principles, techniques, and diagnostic imaging guidance. American Journal of Roentgenology 174(2):323–331, URL http://www.ajronline.org/content/174/2/323.short, http://www.ajronline.org/content/174/2/323.full.pdf+html

Grant MC, Boyd S (2008) Graph implementations for nonsmooth convex programs. In: Blondel V, Boyd S, Kimura H (eds) Recent Advances in Learning and Control, Lecture Notes in Control and Information Sciences, Springer-Verlag Limited, pp 95–110, http://stanford.edu/~boyd/graph_dcp.html

Haase S, Suss P, Schwientek J, Teichert K, Preusser T (2012) Radiofrequency ablation planning: An application of semi-infinite modelling techniques. European Journal of Operational Research 218(3):856–864, DOI 10.1016/j.ejor.2011.12.014, URL http://linkinghub.elsevier.com/retrieve/pii/S0377221711010915

Henriques F (1947) Studies of thermal injury v. the predictability and significance of thermally induced rate processes leading to irreversible epidermal injury. American Journal of Pathology 43:489–502

Henriques F, Moritz A (1947) Studies of thermal injury in the conduction of heat to and through skin and the temperatures attained therein: A theoretical and experimental investigation. American Journal of Pathology 23:531–549

Jiang B, Gao W, Kacher D, Nevo E, Fetics B, Lee TC, Jayender J (2018) Kalman filter-based em-optical sensor fusion for needle deflection estimation. International journal of computer assisted radiology and surgery 13(4):573–583

Livraghi T, Solbiati L, Meloni F, Gazelle S, Halpern E, Goldberg SN (2003) Treatment of focal liver tumors with percutaneous radio-frequency ablation: Complications encountered in a multicenter study. Radiology 226(2):441–451, DOI 10.1148/radiol.2262012198, URL http://dx.doi.org/10.1148/radiol.2262012198, pMID: 12563138, http://dx.doi.org/10.1148/radiol.2262012198

Lloyd S (1982) Least squares quantization in PCM. IEEE transactions on information theory 28(2):129–137

Medtronic (2016) Cool-tip RF Ablation System E Series Electrodes and Accessories. http://www.medtronic.com/covidien/products/ablation-systems/cool-tip-rf-ablation-system-e-series-electrodes, [Online; accessed 04-October-2016]

Moritz A (1947) Studies of thermal injury III. The pathology and pathogenesis of cutaneous burns: An experimental study. American Journal of Pathology 23:915–934

Moritz A, Henriques F (1947) Studies of thermal injury II. The relative importance of time and surface temperature in the causation of cutaneous burns. American Journal of Pathology 23:695–720

Mundeleer L, Wikler D, Leloup T, Lucidi V, Donckier V, Warzée N (2009) Computer-assisted needle positioning for liver tumour radiofrequency ablation. International Journal of Medical Robotics 5(4):458–64, DOI 10.1002/rcs.278, URL http://doi.wiley.com/10.1002/rcs.v5%3A4

Neshat H, Cool DW, Barker K, Gardi L, Kakani N, Fenster A (2013) A 3d ultrasound scanning system for image guided liver interventions. Medical physics 40(11):112,903

NIST (2015) NIST reference on constants units and uncertainity. http://physics.nist.gov/cgi-bin/cuu/Value?r, [Online; accessed 12-January-2013]

O’Rourke AP, Haemmerich D, Prakash P, Converse MC, Mahvi DM, Webster JG (2007) Current status of liver tumor ablation devices. Expert Review of Medical Devices 4(4):523–537, DOI 10.1586/17434440.4.4.523, URL http://dx.doi.org/10.1586/17434440.4.4.523, http://dx.doi.org/10.1586/17434440.4.4.523

Pavlovich C, Walther M, Choyke P, Pautler S, Chang R, Linehan M, Wood B (2002) Percutaneous radio frequency ablation of small renal tumors: initial results. The Journal of urology 167(1):10–15

Pearce J (2009) Relationship between Arrhenius models of thermal damage and the CEM 43 thermal dose. In: Proceedings of SPIE, Energy-based Treatment of Tissue and Assessment V, vol 7181, pp 718,104– 718,104–15, DOI 10.1117/12.807999, URL +http://dx.doi.org/10.1117/12.807999

Pearson AS, Izzo F, Fleming RD, Ellis L, Delrio P, Roh M, Granchi J, Curley S (1999) Intraoperative radiofrequency ablation or cryoablation for hepatic malignancies. The American journal of surgery 178(6):592–598

Peng J, Wei Y (2007) Approximating k-means-type clustering via semidefinite programming. SIAM Journal on Optimization 18(1):186–205

Piccioni F, Poli A, Templeton LC, Templeton TW, Rispoli M, Vetrugno L, Santonastaso D, Valenza F (2019) Anesthesia for percutaneous radiofrequency tumor ablation (prfa): a review of current practice and techniques. Local and regional anesthesia 12:127

Rempp H, Boss A, Helmberger T, Pereira P (2011) The current role of minimally invasive therapies in the management of liver tumors. Abdominal imaging 36(6):635–647

Romeijn H, Dempsey J (2008) Intensity modulated radiation therapy treatment plan optimization. TOP 16(2):215–243, DOI 10.1007/s11750-008-0064-1, URL http://dx.doi.org/10.1007/s11750-008-0064-1

Sapareto S, Dewey W (1984) Thermal dose determination in cancer therapy. International Journal of Radiation Oncology Biology Physics 10(6):787–800, DOI 10.1016/0360-3016(84)90379-1, URL http://www.sciencedirect.com/science/article/pii/0360301684903791

Schwarzmaier HJ, Yaroslavsky AN Ilya V and Yaroslavsky, Fiedler V, Ulrich F, Kahn T (1998) Treatment planning for mri-guided laser-induced interstitial thermotherapy of brain tumors—the role of blood perfusion. Journal of Magnetic Resonance Imaging 8(1):121–127, DOI 10.1002/jmri.1880080124, URL http://dx.doi.org/10.1002/jmri.1880080124

Strauss WA (1992) Partial differential equations: An introduction. New York

Tungjitkusolmun S, Staelin TT, Haemmerich D, Jang-Zern T, Hong C, Webster JG, Lee Jr FT, Mahvi DM, Vorperian VR (2002) Three-dimensional finite-element analyses for radio-frequency hepatic tumor ablation. IEEE Transactions on Biomedical Engineering 49(1):3 –9, DOI 10.1109/10.972834

Villard C, Soler L, Gangi A (2005) Radiofrequency ablation of hepatic tumors: simulation, planning, and contribution of virtual reality and haptics. Computer Methods in Biomechanics and Biomedical Engineering 8(4):215–227, DOI 10.1080/10255840500289988, URL http://www.tandfonline.com/doi/abs/10.1080/10255840500289988

WHO (2012) Media center. http://www.who.int/mediacentre/factsheets/fs297/en/, [Online; accessed 13-December-2012]

Wissler E (1998) Pennes’ 1948 paper revisited. Journal of Applied Physiology 85(1):35–41, URL http://jap.physiology.org/content/85/1/35.abstract, http://jap.physiology.org/content/85/1/35.full.pdf+html

Zhang H, Banovac F, Munuo S, Campos-Nanez E, Abeledo H, Cleary K (2007) Treatment planning and image guidance for radiofrequency ablation of liver tumors. In: Proceedings of SPIE, Medical Imaging 2007: Visualization and Image-Guided Procedures, vol 6509, pp 650,922–650,922–10, DOI 10.1117/12.711489, URL http://dx.doi.org/10.1117/12.711489

